# Getting seniors back on their bicycle; a pretest-posttest case-control study on the improvement of bicycle balance control

**DOI:** 10.1101/2024.04.17.24305755

**Authors:** Eric Maris

## Abstract

This paper quantifies the effectiveness and generalizability of an intervention that aims at restoring bicycle balance control skills in seniors that have quit cycling. The intervention increased the difficulty of the bicycle balance control task in a stepwise fashion, gradually approaching the difficulty of bicycle balance control on the public roads. The intervention involved three components: (1) training on an exercise bicycle, (2) balance control training on a bicycle simulator, and (3) cycling on the public roads with a safe start-and-stop technique that was practiced on the bicycle simulator. The experimental group consisted of 23 community-dwelling senior citizens that had quit cycling (N=19) or that were on the verge of doing so (N=4). The effectiveness of the intervention was evaluated by comparing balance control skill and confidence between a post-and a pre-intervention measurement. To control for a possible spontaneous recovery, the same comparison was also performed in a matched control group that did not participate in the intervention. The intervention produced a very large improvement (Cohen’s d = 1.5, t(16)=6.0, p<.001) in balance control skills and confidence on the public roads. Bicycle balance control skills and confidence can thus be restored over a short period of time. In the Discussion section, I argue that this very large improvement does not rule out the importance of the slower process of acquiring a sufficient lower body strength.

## Introduction

With age, there is a sharp decrease in the percentage of citizens that continues to ride their bicycle. This is well documented in the Netherlands (1), where everyone learns to cycle at a young age, and cycling is the default means of transportation for short distances. In the Netherlands, the percentage of inhabitants that cycles at least several times per month fluctuates around 77% between age 30 and 70 (1).

However, over an average period of 15 years, this percentage decreases sharply to 39% (1).

Using life expectancy statistics (2), one can calculate that the average non-cycling period is 16 years.

The physical health benefits of cycling are well documented (3, 4). For instance, there is a strong inverse relationship between commuter cycling and all-cause mortality (4). Because pedaling force can be easily controlled, cycling is an excellent training stimulus for the improvement of lower body strength and power. In addition, it has a very low impact on the joints, and it offers increased mobility options that are necessary for a meaningful social life.

In this paper, I describe, motivate, and evaluate a rehabilitation intervention focused on balance control (BC) skills that aims at getting senior citizens back on their bicycle. Bicycle BC pertains to two centers of gravity (CoGs) that both must be kept above their respective area of support (AoS): (1) the CoG of the rider-bicycle combination must be kept above the line that connects the two wheel-road contact points, and (2) the CoG of the rider’s upper body must be kept above the saddle (5). To achieve these two objectives, steering (turning the handlebars) is the most important control action (5, 6).

BC can be understood from the perspective of optimal feedback control (OFC) (5, 7-13), the dominant theory in the neural control of movement (see Fig. 1). OFC distinguishes between two determinants of BC skill: (1) the sensory-motor control algorithms implemented in the neuronal networks of our central nervous system (CNS), and (2) strength, power and joint range-of-motion as implemented in the muscles, tendons, and joints (the human part of the mechanical system; see Fig. 1). Our nervous system also has a somatic nervous system (SNS) which sends out motor commands and receives sensory feedback (see Fig. 1).

**Fig. 1.**
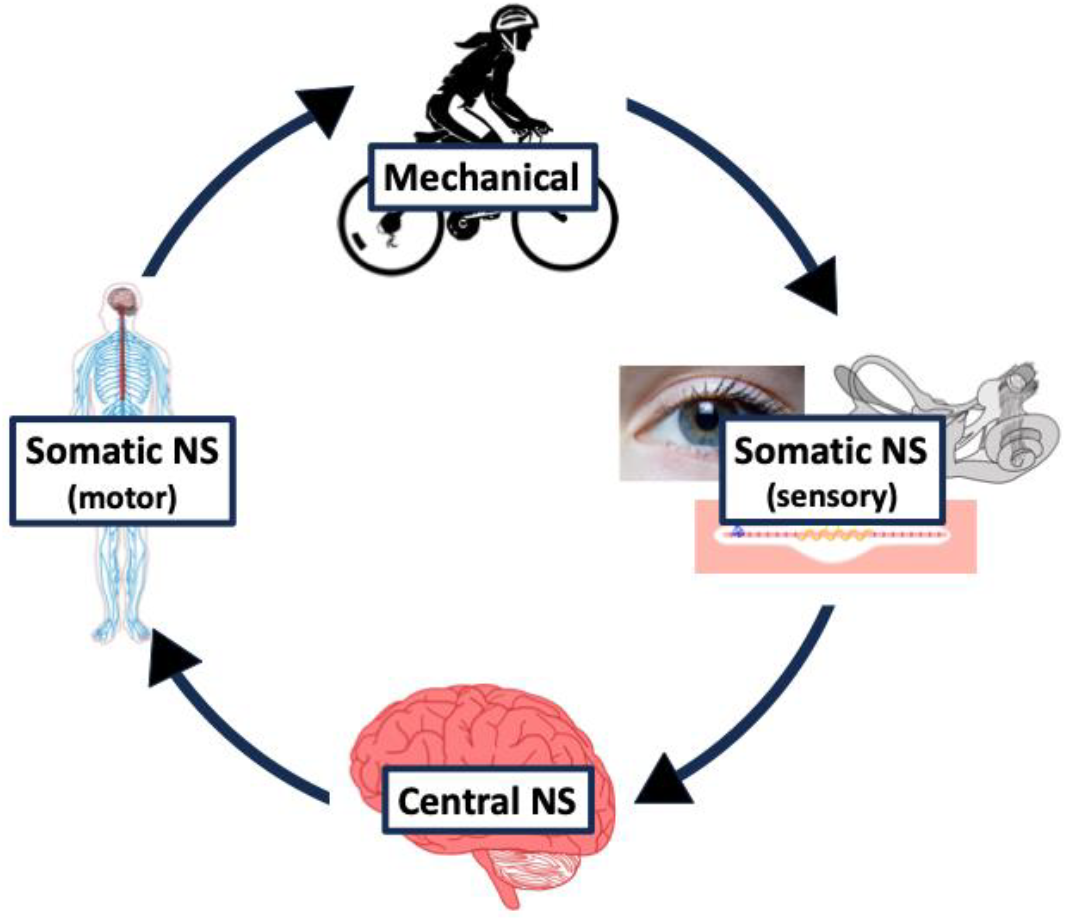
A closed-loop feedback control model for bicycle balance. The model has four components: (1) a sensory system that represents the sensory division of the somatic nervous system (SNS), consisting of the eyes, the inner ear (labyrinth, vestibular organ), the proprioceptive sensory organs (muscle spindles and Golgi tendon organs), and their afferent axons, (2) a computational system that represents the central NS (CNS), (3) a motor output system that represents the sensory division of the SNS, consisting of the efferent axons of the spinal motor neurons, and (4) a mechanical system that represents the non-neuronal part of the human body (extrafusal muscle fibers, skeleton, …) and the bicycle.

OFC is consistent with the fact that the effects of different types of balance training are specific to the balance task that is trained (14, 15). In fact, OFC postulates that, separately for every balance task, the CNS optimizes BC under the constraints/limitations of the mechanical and the SNS (e.g., precision of the sensory feedback, torque and range limits of the mechanical system, motor noise); there is no set of component BC skills that is assembled in different ways according to different sets of task requirements. For a bicycle BC training, this task-specificity would imply that a training effect would not generalize to cycling on the public roads.

For a lost motor skill that must be restored, it is important to know to what extent it depends on (1) plasticity in the CNS (responsible for the mapping of sensory feedback onto control actions), and (2) plasticity in the organs that are responsible for registering feedback and executing actions. Plasticity in the CNS depends on the signaling properties of neurons which can change over a much shorter timescale (hours, days) than the organs that are responsible for sensory registration and motor execution (16-18). To the best of my knowledge, plasticity in the sensory organs has not yet been demonstrated, and plasticity in the muscles with functional consequences (strength/power increase) operates at a timescale of at least weeks.

I designed a bicycle BC training with a double objective: (1) realizing training effects that generalize to bicycle BC on the public roads, (2) dissociating central (CNS-dependent) and muscular contributions to a possible improvement in BC skill. To realize the first objective, the difficulty of the bicycle BC task was increased in a stepwise fashion, gradually approaching the difficulty of bicycle BC on the public roads. And to realize the second objective, the training included sessions on exercise (spinning) bicycles on which the functional threshold power (FTP) could be monitored.

To realize a stepwise increase in the difficulty of the bicycle BC task, it is useful to consider BC skill as a continuum defined by BC challenges, which are perturbations that distort the balance. An important BC challenge are the upward reaction forces that result from pedaling, deform the upper body and perturb the steering. The impact of a BC challenge is modulated by the cycling speed, which is reflected in the fact that it is much easier to balance a bicycle when we are up to speed, as compared to when we are close to a standstill. The modulating role of speed follows from the mechanical properties of the bicycle, which are responsible for self-stabilizing forces that increase with speed (6).

In the bicycle BC training, I make use of the fact that the BC challenge level increases with pedaling force and decreases with speed. To control the BC challenge level, I have developed a bicycle simulator that requires the same type of BC as on the public roads (see Methods). On this simulator, also a safe start- and-stop technique was practiced which allowed the rider to continue with his BC exercises while cycling safely outdoors.

## Study setup and methods

### Participants

Participants were recruited via an article in a local newspaper (De Gelderlander) about the goal of the study (March 31, 2023). This newspaper was distributed over an area with center roughly at the city of Nijmegen, the location of the intervention. The article invited participants that stopped cycling at least six months ago and wanted to get back on their bicycle. A total of 105 candidates replied, which were all invited for an information session on the Radboud University campus in Nijmegen. Based on their responses to a questionnaire, I excluded candidates that still cycled occasionally or suffered from balance-relevant sensory disorders (peripheral neuropathy, labyrinthitis, vestibular neuritis, …). I then selected the 15 participants that lived the closest to the location of the intervention (Radboud University campus). The sole motivation for this selection was the desire to minimize travel time for the participants. As described under “Intervention”, participants had to visit Radboud University campus multiple times.

The study was conducted in accordance with the Declaration of Helsinki, and the protocol was approved by the Ethics Committee of the faculty of Social Sciences (ECSS) of the Radboud University (ECSS-2020-132). All participants gave their written informed consent for inclusion before they participated in the study. This informed consent was obtained in two stages: (1) by completing an online questionnaire in which candidates requested for participation, and (2) by reaffirming this request in a second questionnaire (distributed via email) after the information session.

The intervention started with cycling on a stationary exercise bicycle (spinning) in the second week of June 2023. This part of the intervention was in the Radboud University Sports Center. On June 15, a second newspaper article was published in which the location of the intervention was revealed. This second article was a progress report on the study, and did not ask for participation. However, after this publication, 8 additional candidates spontaneously joined the spinning sessions, and requested to participate in the study, to which I agreed. Four of these additional participants still cycled on rare occasions but considered quitting because of fear of falling.

A total of 23 participants started the BC training on the bicycle simulator, and 17 participants completed it (mean age 75,3 years, age range 66-88 years). From the 6 participants that dropped out, 2 did so for a reason that was related to the BC training: one participant’s legs were so short that she could not touch the ground while sitting on the saddle, and one participant lacked a sufficient force to keep herself balanced on top of a stationary bicycle. Four participants dropped out for reasons that were unrelated to the BC training: a fracture due to a fall at home (two participants), bursitis, and an urgent eye surgery.

The 17 participants that completed the BC training were matched to 17 candidates (the control group) that were not selected for the study because they were living farther away from the location of the intervention as compared to the experimental group. The matching criteria were gender and age.

### Bicycle simulator

The bicycle simulator (19) is shown in Fig. 2. The core elements of the simulator are three rollers with a width of 1.75 m. This is likely to be close to the width of an average Dutch bicycle path because (1) prior to 2022, the advised minimum width was 2 m, and (2) 40 percent of the bicycle paths did not meet this requirement (20). The two front rollers are covered with a conveyer belt that is supported from underneath by a low-friction platform (not visible on Fig. 2). The bicycle’s front wheel rests on the conveyer belt and the rear wheel rests on the rear roller. The rollers’ rotational inertia is close to the linear inertia of the average Dutch rider-bicycle combination, and this ensures that the acceleration response to a pedal stroke feels natural. A torque motor is attached to the axis of the rear roller, and this motor is essential to assist weak participants in the first stages of the training. Finally, the bicycle simulator has a virtual-reality (VR) component that projects an animation of a bicycle lane (not visible on Fig. 2), which is controlled by the cycling speed and the cyclist’s lateral position on the left-right axis of the simulator (as measured using a motion capture system).

**Fig. 2.** Actor riding on the bicycle simulator while being supported by trainers.

In between the front and the rear rollers, there is a platform on which the rider can place his feet and the trainers can stand. The rider has one trainer on every side, supporting him by means of gentle pushes on his shoulders. As the rider’s BC skills improve, the trainers increase their distance from the rider.

The rider wears a safety harness with which he is connected to a support construction above the rider’s head (see Fig. 2). The safety harness does not prevent the rider from falling, but in case he does, his knees will not touch the floor before the suspension becomes active.

### Intervention

The intervention was over a period of 11 weeks. In that period, the participants engaged in (1) spinning, (2) BC training on the bicycle simulator, and (3) cycling on the public roads with a safe start-and-stop technique that was practiced on the bicycle simulator. The three components of the intervention were also administered in this order, with spinning and BC training overlapping in time for the last 8 weeks.

#### Spinning

The first three weeks of the intervention only involved a group spinning training, which was given by the first author and two professional spinning coaches. Participants trained at an individually determined power level, controlled by the one-minute functional threshold power (1minFTP).

One spinning session lasted 45 min., excluding warming-up and cooling-down, but including a 5 min. break in the middle. Every session started with a 3-minute block in which the 1minFTP was assessed. The remainder of the session had the structure of an interval training. The high intensity blocks lasted at most 1 min. and were alternated with low intensity blocks at 70 percent of 1minFTP that lasted at most 2,5 min. Increasing intensity was achieved either by increasing the resistance (and keeping the RPM fixed) or by increasing the RPM (and keeping the resistance fixed).

#### Assessing the 1minFTP

The 1minFTP was used for a double purpose: (1) as a measure of leg power, to investigate its relationship with bicycle BC skill, and (2) to train at an individually determined power level. The 1minFTP is the highest power a participant can maintain for one minute while cycling at 60 revolutions per minute (RPM). The 1minFTP was assessed by having participants monitor their breath after a one-minute effort at some power level: if the participant could say only a few words at a time (a proxy for reaching the ventilatory threshold), the 1minFTP was reached.

Assessing the 1minFTP required keeping a constant high-power output, and this turned out to be very difficult for our participants. First, assessing the 1minFTP required that the participants could operate the bicycle computer that was used to provide feedback about their power output. It took participants two weeks to learn to operate this computer. Second, it took several participants substantially more time to learn to produce a constant power output; initially, all participants lowered their RPM when the resistance was increased above a certain level, thereby preventing a higher power output. Third and last, participants’ performance was highly variable with respect to the maximum power output they endured, and this was both within sessions (no longer following the trainer’s instructions with respect to power output) and between sessions (lowering the 1minFTP at the beginning if one was feeling tired).

#### Bicycle simulator BC training

BC training started in the fourth week of the intervention and was in one of the research labs of the Radboud University (Faculty of Social Sciences). The BC training was individual and was given by the first author and two research assistants. A BC training session lasted approximately 30 min. At the start, participants put on a safety harness and were connected to the support construction. The saddle height was adjusted such that the participant’s feet touched the platform in between the front and the rear rollers. As long as the participant could not balance independently and/or showed signs of fear, the trainers gently pushed their hands against the participant’s shoulders. They gradually released the pressure as the participant’s BC became more independent and he/she gained confidence.

The guiding principle in the BC training was increasing the difficulty in small steps and following the participant’s progressing skill and confidence. Every session started with a BC challenge level at which the participant was comfortable, and that level was slowly increased if the participant agreed. In the first session, the bicycle was brought up to a speed of approximately 12 km./h. by means of a torque motor. In this condition, the participant’s only task was turning the handlebars for balance control. The BC challenge level was increased in four ways: (1) by decreasing the speed, (2) by decreasing the assistive force at the rear roller, (3) by asking participants to accelerate from standstill and to stop at any desired moment, and (4) by asking the participant to tap the support surface with both feet, place them back on the pedals, and to continue pedaling (feet tapping exercise).

#### Cycling outdoors

The participants were instructed to start cycling outdoors on their own bicycles when they could do 10 feet taps in a row without stopping in between, and at a resistance level slightly above the resistance on the public roads. The participants were instructed to use the safe start-and-stop technique (feet touching the road surface) that was practiced on the bicycle simulator, to start cycling in an environment without other road users (a park, a parking lot), and to gradually move to more busy environments. A few participants informed me that they had started cycling outdoors before I had instructed them to do so. Those participants were also instructed to use the safe start-and-stop technique, to start cycling in an environment without other road users, and to gradually move to more busy environments.

### Causal effect estimation in a pretest-posttest design

A randomized experiment is often considered the only possible way to obtain an unbiased estimate of a causal effect. However, according to statistical theory this is not the only way (21). In fact, if spontaneous recovery can be ruled out (also denoted as the assumption of temporal stability (21)), a pretest-posttest design also allows for such an unbiased estimate. The argument involves that if, in the population of interest, the average of some outcome variable (e.g., bicycle BC skill) does not change spontaneously, then the average post-minus pre-intervention change in this outcome variable is an unbiased estimate of the treatment’s causal effect. For bicycle BC skill in our population of interest (senior citizens), it is unlikely that this skill spontaneously returns (i.e., without a deliberate intervention) after it has been lost. Of course, numerous other outcome variables do change spontaneously on average (body temperature in a population that has the flu, feelings of depression in a population that visits a psychotherapist, walking BC skills in healthy young adults that had bed rest for a week, …). To remove all doubts about a possible spontaneous recovery, I included a control group that did not receive the intervention, but in which the change in the outcome variable was assessed over the same time interval as in the experimental group (see Participants).

The effect of the intervention was then evaluated statistically using a dependent samples t-test that compared the change in the outcome variable between the matched experimental and the control group participants.

### Outcome variable

The outcome variable was obtained from a questionnaire that evaluated participants’ bicycle BC skills and confidence while riding on the public roads. To maximize the validity of this questionnaire, the questions were formulated in terms of observable criteria. The introduction to the questionnaire was the following:

> Three environments will be described, and you must indicate whether you rode your bicycle in that environment in the past two weeks. By “riding your bicycle”, we mean starting from a standstill and stopping at a desired location. For every environment, you must indicate what applies to you: (1) I did not ride my bicycle here, (2) I rode my bicycle here, but I try to avoid it, and (3) I rode my bicycle here without problems. The three environments are the following: (1) without traffic (e.g., a park, a parking lot), (2) with little traffic (e.g., small roads in an outside area, a bicycle lane on a quiet moment), and (3) in busy traffic (e.g., in the city center during traffic hours, around a busy marketplace).

The answers to every question/environment were converted into a score by assigning 0, 1, and 2 points to the respective options. Summing these scores over the three questions/environments results in a number between 0 and 6, which is the outcome variable.

In the experimental group, this questionnaire was administered twice, the first time in the beginning of the intervention and the second time six weeks after the end of the intervention. The questionnaire was sent as an attachment to an email, and participants were called on their phone if they had not replied after one week. For some participants, it was necessary to clarify some aspects of the questions. In the control group, the questionnaire was administered only once and at around the same time that the second questionnaire was sent to the experimental group. None of the control group participants was riding their bicycle at the start of the intervention and they were asked whether and how this had changed in the meantime.

## Results

I conducted a pretest-posttest case-control study to evaluate the effectiveness of an intervention aimed at restoring bicycle BC skill. The experimental group consisted of 23 participants, of which 17 completed the intervention (see Methods).

The intervention lasted 11 weeks and involved three components: (1) cycling on a stationary exercise bicycle (spinning), (2) BC training on a bicycle simulator, and (3) cycling on the public roads with a safe start-and-stop technique that was practiced on the bicycle simulator (see Methods). The effectiveness of the intervention was evaluated in terms of the participant’s skill/confidence on his own bicycle when cycling outdoors (see Methods). In the experimental group, this outcome measure was assessed twice, first in the beginning of the intervention, and then six weeks after the intervention. To correct for a possible spontaneous recovery of cycling skill/confidence, I used a matched control group that did not receive the intervention, but in which the change in cycling skill/confidence over the same time interval was assessed (see Methods).

The improvement in cycling skill/confidence was significantly larger in the experimental group (M=3.1, SD=2.1) compared to a matched control group (M=0, SD=0), t(16)=6.0, p<.001, 95% CI [2.0 4.1]. Note that none of the control group participants had a spontaneous recovery of their cycling skill/confidence. The raw outcome measures of the experimental group are shown in Fig. 3. The effect size (Cohen’s d) was 1.5, which is between a very large and a huge effect (22). This effect was obtained with an average of 4.8 simulator BC training sessions of 30 min. each.

**Fig. 3.**
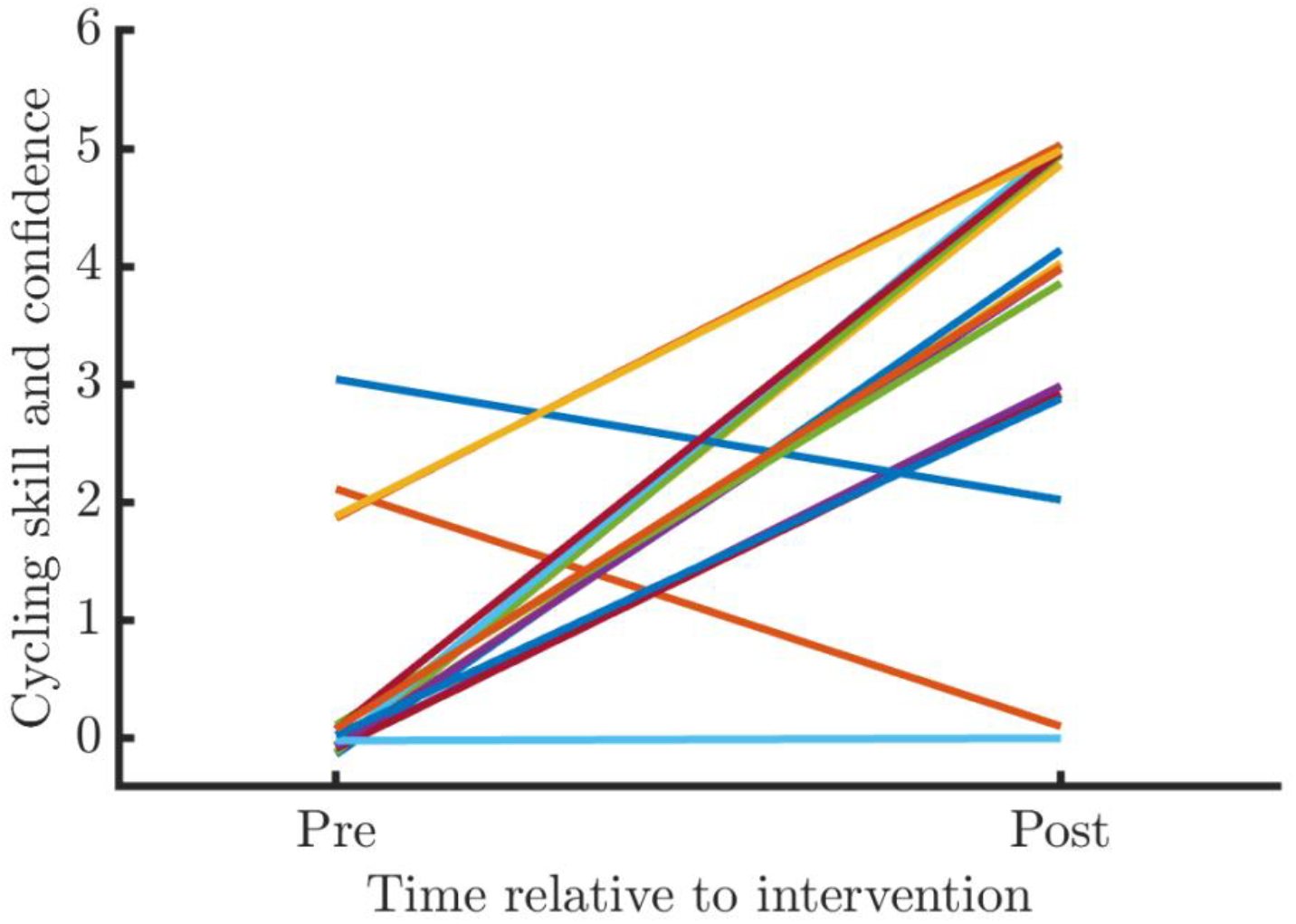
Raw outcome measures of the experimental group (a small random jitter was added to see overlapping lines)

It was my original plan to dissociate central (CNS-dependent) and muscular contributions to a possible improvement in BC skill. For that, it was necessary to quantify the increase in maximum power output on the spinning bicycle, for which I used the one-minute functional threshold power (1minFTP), which was assessed at the beginning of every spinning session. However, keeping a constant high-power output (a requirement for a reliable FTP assessment) turned out to be very difficult for our participants and required several weeks of training (see Methods). I conclude that I failed in obtaining a reliable quantification of the increase in maximum leg power increase in this group of participants.

However, it is useful to report the mean 1minFTP on which participants converged after learning to produce a constant force output. This mean 1minFTP was 93.6 Watt (sd=17.2). FTP is usually expressed relative to bodyweight, and in this participant group the average 1 minute power-to-weight ratio was 1.2 Watt/kg (sd=0.3). For reference, it is useful to compare this value with published 1 minute power-to-weight ratios for untrained amateur cyclists (23) (defined as the 5-th percentile of the population of amateur cyclists): 3.9 for males and 3.3 for females. Thus, the participants in this study had an average 1minute power-to-weight ratio that is approximately three times smaller than that of an untrained cyclist.

## Discussion and conclusions

A very large improvement in senior cyclists’ BC skills and confidence can be realized by means of a short intervention (11 weeks), even if that person has quit cycling for a long time. This is remarkable given the task-specificity of the effects of different types of balance training (14, 15), and is most likely due to the high similarity between the BC training on the simulator and cycling outdoors.

### Limitations

This study did not expose participants to some important BC challenges such as an uneven road surface (e.g., curbstones, potholes) and sharp turns, and it is unknown whether they can deal with them successfully. It is very likely that several BC challenges require a sufficient lower body strength to prevent perturbations from affecting the steering movements. For example, (1) standing on the pedals while covering an obstacle, and (2) choosing an appropriate lean angle before entering a turn.

Cycling safely not only involves BC (staying upright) but also navigation (avoiding obstacles and/or following a track), which requires additional skills such as selective attention, anticipation, and planning. These have not been investigated.

In the absence of an existing objective measure of BC skills on the public roads, I used a self-report measure with questions that were formulated in terms of observable criteria. Although suboptimal in general, such a self-report measure is to be preferred over the objective measures that were collected as a part of the BC training (e.g., the assistive force required to accelerate from standstill, the number of times the rider could tap his/her feet on the support surface). In fact, there is no information about these objective measures’ predictive value for BC skills on the public roads.

From the perspective of OFC, it is tempting to conclude that the improvement is mainly due to the fast plasticity in the CNS, and not to the comparatively slow plasticity in the muscles. However, to make a strong case for this claim, it is essential to have a reliable measure of maximum power output, and this turned out to be very difficult. The existing literature on the relation between (1) muscle strength/power, and (2) the incidence of falls and balance task performance is inconsistent, with one systematic review (focusing on falls) (24) arguing for a strong relation and another (focusing on balance task performance) (25) for a very weak one. This inconsistency is probably related to the fact that, in the population of interest, falls cannot be predicted from the performance on balance tasks (26).

For several reasons, the relation between muscle strength/power, balance control, and falls, is a complicated one. First, many studies have been conducted in homogeneous groups (institutionalized participants, a small age range, …) and this restriction-of-range results in an underestimation of the correlation in the heterogeneous/unselected population. Crucially, restriction-of-range also operates at the individual level because most people are aware of the range of BC challenges that their BC skill allows, and they try to stay within this range. This restriction-of-range at the individual level reduces the correlation between, on the one hand, muscle strength/power and BC skills, and on the other hand, the incidence of falls. Experimental studies in which falls are elicited do not suffer from this restriction-of-range at the individual level, and this methodology has been applied successfully to investigate tripping-induced falls during walking (27, 28). One such study found a convincing correlation between leg strength and the ability to recover from tripping(28).

Second, it is very likely that there are large differences between muscle groups and their associated joint ranges-of-motion with respect to their relevance for BC, and that this differential relevance depends on the BC task. BC during walking, such as the ability to recover from tripping, must depend on leg strength because the leg muscles are used to bring the body CoG back over its AoS. For bicycle BC, on the other hand, different muscle groups are used to control the combined rider-bicycle and the upper body CoG: (1) the arm muscles (which control the handlebars) bring the combined rider-bicycle CoG over its LoS, and (2) the core muscles in the trunk and the hip bring the upper body CoG over its AoS, the saddle. Thus, the relevance of a particular muscle group for balance control likely depends on the specific BC task.

Third and last, BC not only depends on the muscles for bringing the CoG above the AoS (the muscles’ motor role) but also for informing the CNS about the position of CoG relative to the AoS, which depends on the muscle spindles (the muscles’ sensory feedback role) (13, 29). To be useful, muscle spindle output must reflect the tension in the force-producing fibers, but its precise contribution to the loss of BC is unknown.

## Conclusion

I have demonstrated that it is possible to realize a very large improvement in a senior cyclist’s BC skills and confidence over a short period (11 weeks). This is most likely due to the high similarity between the BC training and cycling outdoors, but it does not rule out the importance of the slower process of building (and losing) muscular strength/power.

## Data Availability

All data produced in the present study are available upon reasonable request to the authors

## Author’s contributions

All authors contributed equally to the manuscript and read and approved the final version of the manuscript.

## Acknowledgments

The author would like to thank Mauritz Kleyn and Mirna Linden for their assistance with the spinning training and Maartje Floris and Loes van Leeuwen for their assistance with the bicycle simulator training and the data collection.

## References

1. Mobycon. Research facts and numbers ‘Cycling for everyone’ [Onderzoek feiten en cijfers ‘Fietsen voor Iedereen’]. Delft: Mobycon, commissioned by the Ministery of Infrastructure and Water Management.; 2022.

2. VZinfo.nl. Bilthoven: RIVM; 2024 [Available from: https://www.vzinfo.nl/levensverwachting/leeftijd-en-geslacht#Levensverwachting.

3. De Hartog JJ, Boogaard H, Nijland H, Hoek G. Do the health benefits of cycling outweigh the risks? Environmental health perspectives. 2010;118(8):1109–16.

4. Oja P, Titze S, Bauman A, De Geus B, Krenn P, Reger-Nash B, et al. Health benefits of cycling: a systematic review. Scandinavian journal of medicine & science in sports. 2011;21(4):496–509.

5. Maris E. A bicycle can be balanced by stochastic optimal feedback control but only with accurate speed estimates. PLoS one. 2023;18(2):e0278961.

6. Meijaard JP, Papadopoulos JM, Ruina A, Schwab AL. Linearized dynamics equations for the balance and steer of a bicycle: a benchmark and review. Proceedings of the Royal Society of London A: Mathematical, Physical and Engineering Sciences. 2007;463(2084):1955–82.

7. Åström KJ. Introduction to stochastic control theory: Courier Corporation; 2012.

8. Todorov E. Optimality principles in sensorimotor control. Nature neuroscience. 2004;7(9):907–15.

9. Todorov E, Jordan MI. Optimal feedback control as a theory of motor coordination. Nature neuroscience. 2002;5(11):1226–35.

10. Wolpert DM. Computational approaches to motor control. Trends in cognitive sciences. 1997;1(6):209–16.

11. Scott SH. Optimal feedback control and the neural basis of volitional motor control. Nature Reviews Neuroscience. 2004;5(7):532–45.

12. Wolpert DM, Ghahramani Z, Jordan MI. An internal model for sensorimotor integration. Science. 1995;269(5232):1880–2.

13. Maris E. An internal sensory model allows for balance control based on non-actionable proprioceptive feedback. arXiv. 2024.

14. Giboin L-S, Gruber M, Kramer A. Task-specificity of balance training. Human movement science. 2015;44:22–31.

15. Kümmel J, Kramer A, Giboin L-S, Gruber M. Specificity of balance training in healthy individuals: a systematic review and meta-analysis. Sports Medicine. 2016;46:1261–71.

16. Debanne D, Inglebert Y, Russier M. Plasticity of intrinsic neuronal excitability. Current opinion in neurobiology. 2019;54:73–82.

17. Magee JC, Grienberger C. Synaptic plasticity forms and functions. Annual review of neuroscience. 2020;43:95–117.

18. Mozzachiodi R, Byrne JH. More than synaptic plasticity: role of nonsynaptic plasticity in learning and memory. Trends in neurosciences. 2010;33(1):17–26.

19. Maris E, inventorBicycle simulator [Patent]. The Netherlands2022 01 June 2022.

20. CROW-Fietsberaad. Updated recommendations for cycle path width 2022 [Geactualiseerde aanbevelingen voor de breedte van fietspaden 2022]. In: Waterstaat] MoIaWMMvIe, editor. Ede: CROW-Fietsberaad; 2022.

21. Holland PW. Statistics and causal inference. Journal of the American statistical Association. 1986;81(396):945–60.

22. Sawilowsky SS. New effect size rules of thumb. Journal of modern applied statistical methods. 2009;8(2):26.

23. Johnstone D. How does your cycling power output compare? 2018 [Available from: https://www.cyclinganalytics.com/blog/2018/06/how-does-your-cycling-power-output-compare.

24. Moreland JD, Richardson JA, Goldsmith CH, Clase CM. Muscle weakness and falls in older adults: a systematic review and meta-analysis. Journal of the American Geriatrics Society. 2004;52(7):1121–9.

25. Muehlbauer T, Gollhofer A, Granacher U. Associations between measures of balance and lower-extremity muscle strength/power in healthy individuals across the lifespan: a systematic review and meta-analysis. Sports medicine. 2015;45:1671–92.

26. Boulgarides LK, McGinty SM, Willett JA, Barnes CW. Use of clinical and impairment-based tests to predict falls by community-dwelling older adults. Physical therapy. 2003;83(4):328–39.

27. Pijnappels M, Bobbert MF, van Dieën JH. Push-off reactions in recovery after tripping discriminate young subjects, older non-fallers and older fallers. Gait & posture. 2005;21(4):388–94.

28. Pijnappels M, Van der Burg J, Reeves ND, van Dieën JH. Identification of elderly fallers by muscle strength measures. European journal of applied physiology. 2008;102:585–92.

29. Proske U, Gandevia SC. The proprioceptive senses: their roles in signaling body shape, body position and movement, and muscle force. Physiological reviews. 2012.

